# New Approach Methodologies in Crohn’s Disease Link Molecular Disease Subtypes to Clinical Outcomes

**DOI:** 10.1101/2025.04.01.25325058

**Authors:** Harrison M. Penrose, Saptarshi Sinha, Courtney Tindle, Kameron Zablan, Helen N. Le, Jennifer Neill, Pradipta Ghosh, Brigid S. Boland

**Author notes:** **Senior corresponding authors:** Pradipta Ghosh, M.D.;, Brigid S. Boland, M.D. **Lead contacts:** Pradipta Ghosh, M.D.;, Brigid S. Boland, M.D.

## Abstract

Current clinical decision-making is hindered by the absence of predictive preclinical models that faithfully bridge molecular diversity to patient outcomes. Here, we apply the principle of abstraction—deriving essential features from human tissues to build next-generation new approach methodologies (**NAMs**) that transform patient-derived organoids (**PDOs**) into predictive vehicles for Crohn’s disease (CD). From our living biobank of adult stem cell–derived colonic PDOs, we previously defined two molecular CD subtypes: Immune-Deficient Infectious CD (*IDICD*) and Stress and Senescence-Induced Fibrostenotic CD (*S2FCD*), each defined by unique genomic, transcriptomic, and functional profiles with matched therapeutic vulnerabilities. In this study, we prospectively anchored PDO-derived molecular phenotypes to real-world clinical outcomes, revealing that *S2FCD* maps to baseline and progressive colonic disease activity, whereas *IDICD* tracks with prior ileocecal surgery, penetrating disease behavior, as well as baseline and progressive ileal disease activity. By abstracting NAMs from human tissues and cycling insights between small-‘n’ organoids and Phase 3-sized datasets, this framework recasts PDOs as dynamic, predictive platforms that capture the past, present, and future of disease behavior. Beyond oncology, this work establishes PDOs as vehicles for prospective clinical trial-like studies in inflammatory diseases and highlights colonic immune dysfunction as a potential driver of ileal CD.

**GRAPHICAL ABSTRACT:** 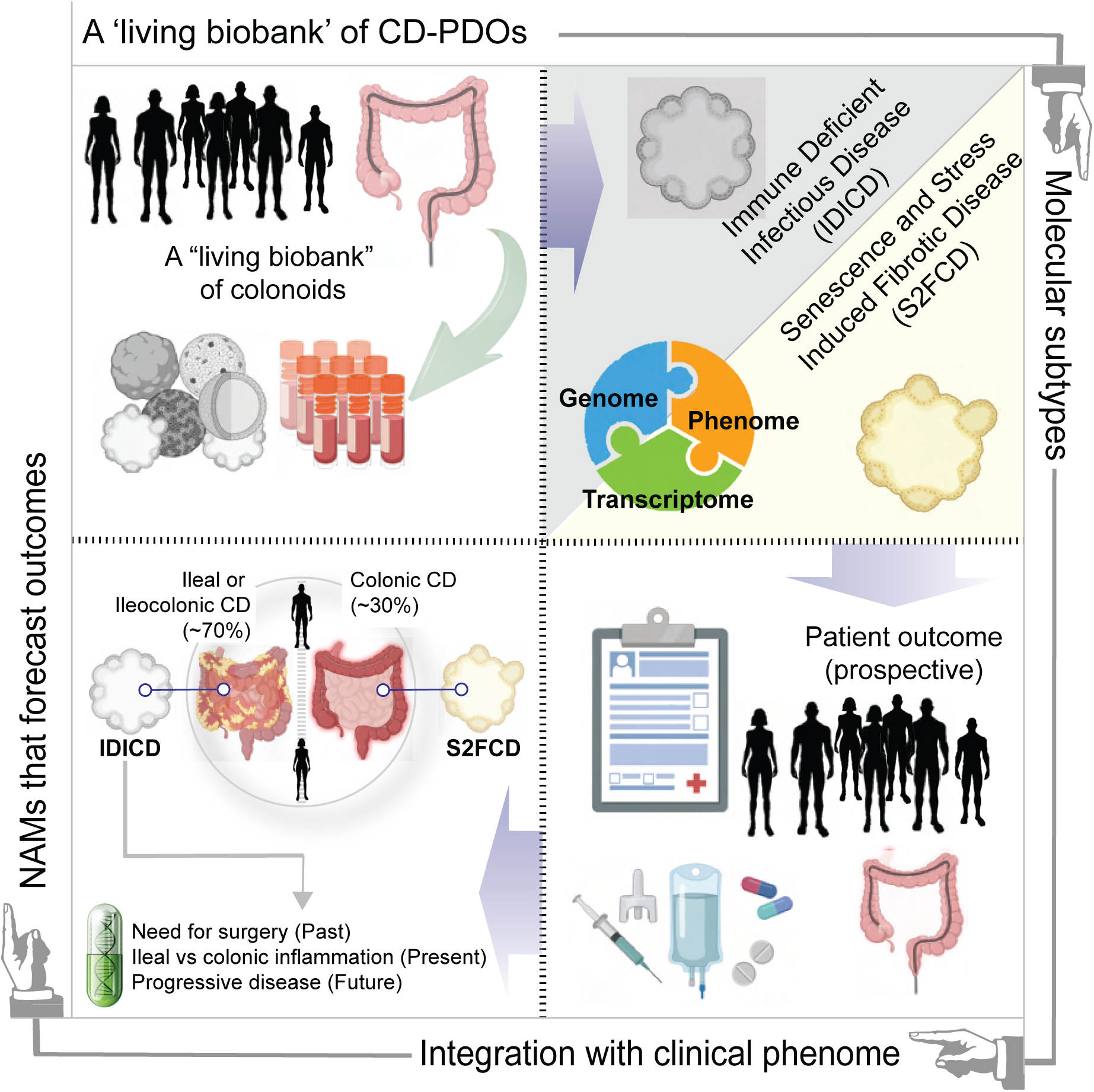

## INTRODUCTION

Inflammatory Bowel Diseases (IBD), encompassing Crohn’s disease (CD) and ulcerative colitis (UC), are chronic inflammatory disorders of the gastrointestinal tract with no perfect preclinical model or cure^1^. Disease pathogenesis and progression are driven by remitting, relapsing periods of inflammation involving dysregulated interactions between the host genome, microbiome, gut immune system, and environmental triggers^2^. CD, in particular, is a remarkably heterogeneous disease, exhibiting highly variable clinical presentations and disease trajectories that remain difficult to predict. While host genetics have partly explained certain disease characteristics (i.e. anatomic location), they fail to account for variations in disease behavior, therapeutic response, environmental susceptibility, and complications, highlighting the need for personalized treatment strategies^3^.

Accurately modeling this heterogeneity has proven especially challenging. Murine systems, while invaluable, cannot fully recapitulate the layered human biology underlying IBD^4^. This gap underscores the growing need for human-derived new approach methodologies (**NAMs**) that are both faithful and scalable, serving as platforms for mechanistic discovery and outcome-linked prediction models.

Patient-derived organoids (PDOs) exemplify this promise as they preserve patient-specific genetics, epigenetics, and disease heterogeneity, while allowing experimental manipulation under controlled conditions. Recent studies have shown PDOs can capture key epithelial dysfunctions characteristic of IBD, but most efforts have been constrained by limited cohort sizes and a lack of real-world clinical anchoring^5,6^.

To address these limitations, we previously developed a living biobank of adult stem cell (AdSC) derived organoids isolated exclusively from fresh **colonic** mucosal biopsies at the UC San Diego IBD Biobank^7^. The living biobank design enables continuous expansion and linkage of PDO phenotypes to real-world patient outcomes over time, elevating PDOs beyond descriptive models into prospective clinical tools. We prioritized the colon over the ileum as it is affected in approximately 60–70% of patients with Crohn’s disease^8^. Additionally, we used AdSCs instead of induced pluripotent stem cells (iPSCs), as AdSCs retain tissue-specific identity and epigenetic imprinting shaped by the exposome of their disease-afflicted organ of origin^9–14^. In our prior work, we analyzed prospectively biobanked colonic PDOs from 24 CD patients, spanning the full spectrum of clinical phenotypes. This design mirrored the real-world heterogeneity of patients treated at tertiary care centers^7^. Deep phenotyping coupled with multi-omics revealed that all PDOs reproducibly collapsed into **two molecular subtypes**: *Immune-Deficient Infectious CD (IDICD)*, marked by impaired pathogen clearance, paradoxical immunosuppression, and CD-risk alleles (NOD2, ATG16L1), and (ii) *Stress and Senescence-Induced Fibrostenotic CD (S2FCD)*^7^, defined by epithelial senescence, profibrotic signaling, and YAP1–IL-18 pathway mutations. These subtypes showed distinct morphologies, transcriptomes, and non-overlapping treatment responses independent of anatomical location (i.e., right vs left colon), inflammation status (i.e., inflamed vs uninflamed), or disease activity state (i.e., active vs inactive CD). These molecular distinctions also carried therapeutic consequences: each subtype required its own targeted intervention, without crossover efficacy, underscoring the need for subtype-specific interventions.

Whereas our prior study focused on molecular classification, the present work advances PDOs as prospective translational tools by linking molecular subtypes to clinical outcomes. By leveraging NAMs anchored in a living biobank, we demonstrate that PDOs can capture the full spectrum of human CD trajectories—from molecular subtype to real-world clinical phenotypes. This approach transforms PDOs into predictive vehicles for precision medicine, bridging discovery and translation, and reframes IBD research through the disciplined abstraction of human biology that is both scalable and clinically meaningful.

## RESULTS

In prior work, we classified CD PDOs into two molecular subtypes–*IDICD* and *S2FCD–*based on multi-omic phenotyping. Although each subtype was distinct from healthy PDOs, direct head-to-head comparisons between *IDICD* and *S2FCD* were not performed. Here, we leveraged the NAMs framework of our living biobank to directly compare these subtypes, treating them as abstracted, human-derived models for prospective outcome prediction.

Differential gene expression revealed clear subtype-specific transcriptional programs (**Fig 1A, B**; see **Table S1** for all DEGs). *IDICD* was marked by upregulation of genes involved in inflammation-immune regulation (*IL33, F2RL1*), sulfation metabolism and detoxification (*SULT1A4, PAPSS2*), and epithelial-stem cell maintenance (*KLF4*). In contrast, *S2FCD* exhibited enrichment of Wnt pathway and developmental genes (*WNT6, NOTUM*), epithelial barrier integrity markers (*LCN2, CDH11*), and immune activation genes (*CD40, TNFRSF6B*) (**Fig 1A**). KEGG (Kyoto Encyclopedia of Genes and Genomes) analysis revealed distinct pathway enrichment for each molecular subtype (**Fig 1B**). *S2FCD* was enriched for pathways consistent with senescence-associated signaling, while *IDICD* unexpectedly showed signatures of retinol and bile acid metabolism, processes typically confined to the ileum^15,16^. Together these findings indicate that *IDICD* and *S2FCD* represent biologically distinct epithelial subtypes of CD, each defined by unique transcriptional programs and signaling pathways. Importantly, this demonstrates that PDOs—abstracted from tissue but anchored in human biology—can recapitulate disease-relevant diversity, thereby functioning as NAMs capable of mapping molecular subtypes to clinical trajectories.

**Figure 1.**
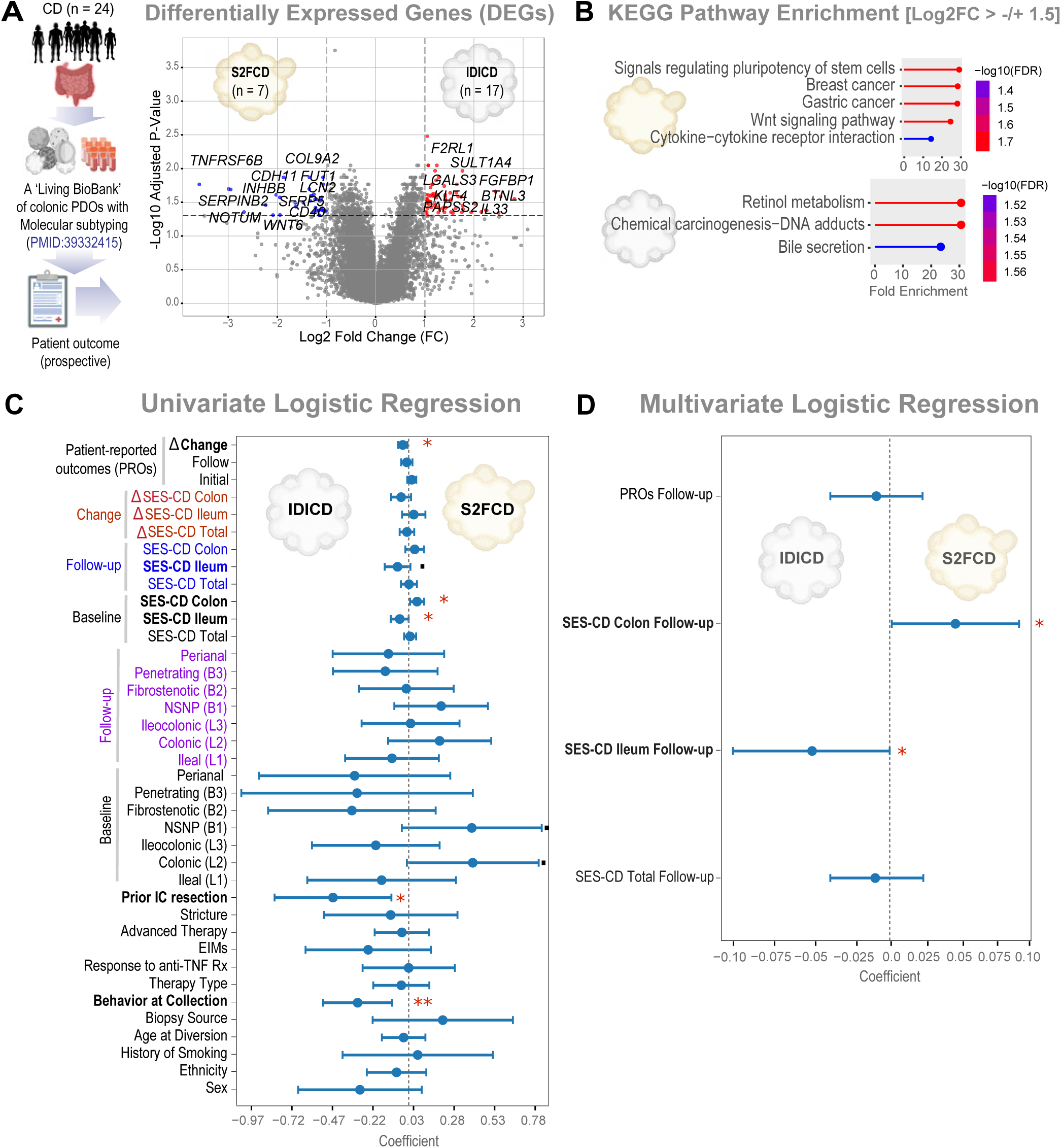
Colonic PDOs hint at distinct pathophysiology and outcomes in ileal versus colonic CD. **(A)** *Left*: Workflow in creating a ‘living biobank’ of colonic PDOs with prospective follow-up on disease outcomes. *Right*: Volcano plot depicting DEGs from RNA-seq in *IDICD* vs. *S2FCD*, where the interrupted line marks log2FC = 1.0. For clarity, gene symbols are shown only for the most interesting genes relevant to CD/IBD. **(B)** Lollipop Bar Plots of KEGG pathways associated with DEGs [log2FC > −/+ 1.5] in A. **(C)** Univariate analysis models of *IDICD* vs. *S2FCD* subtypes as a linear combination of all variables in a CD cohort. The coefficient of each variable (at the center) with 95% CI (as error bars) and the p values are illustrated. The p-value for each term tests the null hypothesis that the coefficient is equal to zero (no effect). ×p ≤ 0.1; *p ≤ 0.05; **p ≤ 0. 01. Bold = significant co-variates. SES-CD, Simple endoscopic score for Crohn’s Disease; IC, ileocecal; EIM, extraintestinal manifestations. Montreal classification for disease behavior: L1 – ileal, L2 – colonic, L3 – ileocolonic; B1 – non-stricturing, non-penetrating; B2 – stricturing; B3 – penetrating. Δ refers to the delta change between baseline and follow-up SES-CD or PRO scores over a prospective five-year follow-up period. Behavior at collection refers to Montreal classification at time of biopsy; here, IDICD is linked to penetrating disease (B3). **(D)** Multivariate analysis of *IDICD* vs. *S2FCD* subtypes as a linear combination of future objective clinical outcomes from the same CD cohort over the prospective five-year follow-up period. The coefficient of each variable (at the center) with 95% CI (as error bars) and the p values are illustrated. The p-value for each term tests the null hypothesis that the coefficient is equal to zero (no effect). *p ≤ 0.05. Bold = significant co-variates. SES-CD, Simple endoscopic score for Crohn’s Disease; PRO, Patient-reported outcomes.

To assess translational relevance, we next asked whether these molecular distinctions forecasted clinical outcomes. Using univariate logistic regression^17^ (UV) across a comprehensive clinical dataset (**Table S2**), we found that *IDICD* subtype showed strong, significant associations with prior ileocecal resection, penetrating disease behavior (B3) at biopsy collection at which time organoids were derived, ileal disease activity (via SES-CD) at baseline, as well as lack of symptom improvement (via increase in PROs) over the five-year follow-up period (**Fig 1C**). In contrast, *S2FCD* correlated with colonic disease activity at baseline (as measured by colonic SES-CD) (**Fig 1C**). Three other variables trended towards significance (p<0.1, indicative of a <10% probability that the observed relationship is due to random chance): *IDICD* was associated with ileal disease activity during follow-up endoscopy and *S2FCD* was associated with a non-stricturing, mucosa-restricted colonic CD phenotype (non-stricturing, non-penetrating; NSNP CD) predominantly isolated to the colon (p<0.1). These findings suggest that while *IDICD* is associated with predominantly ileal disease, *S2FCD* primarily presents as colonic inflammation. Using multivariate (MV) logistic regression, we evaluated the effect of potential confounders to assess whether clinical correlates of disease progression persisted. MV analysis revealed that the association with prospective, future disease activity measures remained robust: ileal disease activity at follow-up (via SES-CD) was significantly associated with *IDICD* subtype, while colonic disease activity at follow-up was significantly associated with *S2FCD* subtype (**Fig 1D**). Together, these results demonstrate that PDO-derived molecular subtypes are independently predictive of future disease trajectories, with *IDICD* molecular subtypes forecasting future ileal disease activity and *S2FCD* predicting subsequent colon-predominant inflammation. Notably, our cohort composition (25% ileal, 46% ileocolonic, 29% colonic) mirrors real-world distributions^8^, supporting the generalizability of these associations. Taken together, these results establish colonic PDOs not just as descriptive models, but as predictive NAMs that link epithelial subtypes to patient outcomes. This demonstrates how living biobanks, when abstracted into reproducible frameworks, can prospectively model the past, present, and future of disease behavior—providing the missing bridge between molecular subtyping and clinical translation.

## DISCUSSION

The major discovery we report here is that two distinct molecular subtypes of colon-derived CD PDOs have equally distinct and meaningful clinical correlates: *IDICD* tracks with baseline and future ileal disease, whereas *S2FCD* aligns with baseline and future colonic disease. Notably, *IDICD* PDOs not only captured ileal disease activity but also reflected a continuum of progression, linking prior ileocecal resection with future disease activity. These correlations elevate PDOs from static descriptors to dynamic, NAM-like platforms: abstracted from human tissue, yet prospectively tied to outcomes in the clinic.

This distinction is critical for therapeutic translation. Prior work demonstrated that each subtype carries unique, reversible defects requiring matched interventions^7^. The ability to track clinical correlates in PDOs during therapeutic reversal means these models can do more than reveal biology; they can serve as prospective vehicles to test whether correcting molecular defects reshapes clinical phenotypes. In this way, CD PDOs achieve the same translational status already attributed to cancer PDOs, validated as predictive precision tools in oncology^18–23^. Here, they show the capacity to recapitulate evolutionary disease trajectories in IBD, enabling personalized risk stratification and mechanism-informed therapeutic strategies.

Two additional insights emerge. First, colonic epithelial defects—particularly innate immune dysfunction in *IDICD*—may serve as surrogates for ileal disease. This suggests systemic and local mechanisms: systemic cytokine or hormonal signals influencing Peyer’s patches, and local retrograde microbial migration in the setting of dysbiosis or obstruction. Second, our PDO-based stratification reinforces the division of CD into ileal-dominant and colonic phenotypes, converging with genetic, epigenetic, microbial, and transcriptomic evidence (**Fig 2D**). For example, transcriptomic analyses of colonic mucosal biopsies have revealed gene expression patterns that distinguish ileal-dominant from colonic CD, with epigenomic regulation differing by disease location^24^. Multi-omics analyses of stool samples have identified microbial, metabolic, and immune signatures—such as bile acid–rich environments and immunoproteomic profiles— that differ between ileal and colonic CD^25,26^. At the cellular level, collagen-producing myofibroblast populations are enriched in ileal disease^27^, while Th1/Th17 populations show variable enrichment depending on disease location^28^. Although not specifically derived from colonic mucosa, genetic studies have supported this subclassification: *NOD2* and *ATG16L1* SNPs have been linked to ileal CD, while *MHC* SNPs correlate with colonic disease^29^. This is also supported through epidemiologic studies showing that smoking is associated with ileal CD, whereas colonic CD is more prevalent in females^30–32^. While previous studies have approached this distinction through genetic, microbial, and transcriptomic lenses, our organoid-based classification provides a functional, epithelial–driven framework that complements and strengthens this evolving model of disease subtypes in CD.

**Figure 2.**
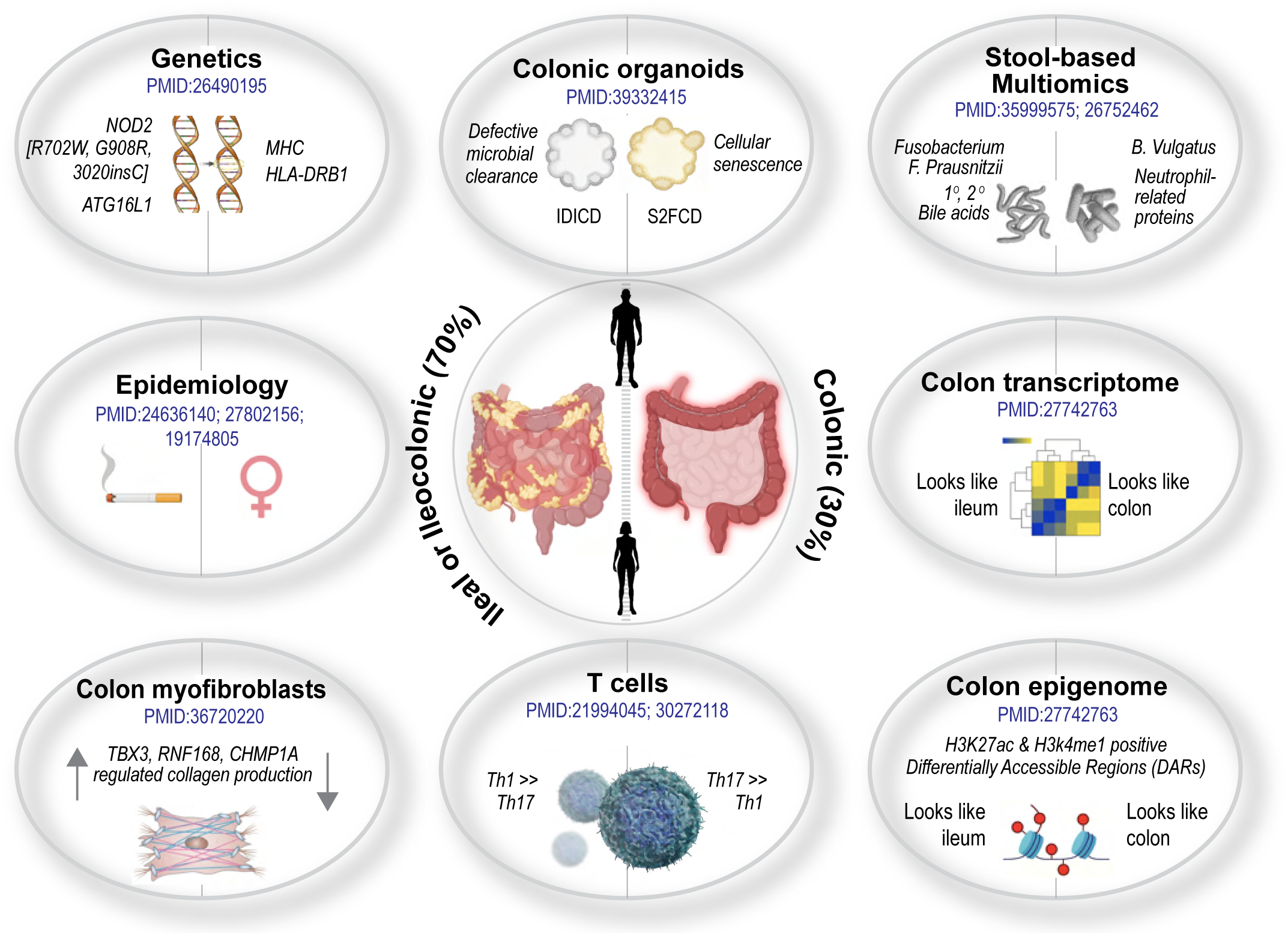
Colonic tissue-focused studies highlight anatomic stratification of CD into ileal vs colonic subtypes. Schematic summarizes a focused, pertinent literature review that converges on the notion that all CDs, regardless of how they were evaluated across scales—genetic, epigenetic, cellular, exposome (stool composition), or clinical/epidemiologic correlates—appear to be broadly classifiable as ileal and/or ileocolonic vs. isolated colonic disease. Colon-focused analyses that consistently arrive at that conclusion, even when uninvolved, are highlighted. Manuscripts were selected based on their analysis of disease location and molecular or clinical phenotyping, with emphasis on studies evaluating colonic tissue.

These findings may carry direct therapeutic implications. Limited efficacy of conventional immunosuppressants in ileal CD, which is more prone to complications^33,34^, may be explained by our prior findings demonstrating the need for rectifying microbial clearance pathways such as dual PPARα/γ agonists to restore epithelial defense mechanisms in *IDICD*^7,35,36^. Given that *IDICD* PDOs show insufficient cytokine response in the setting of infection^7^, it could explain why ileal CD can be less responsive to a broad range of anti-cytokine therapies, including anti-TNFα, anti-IL12/23, and likely JAK inhibitors, all of which often demonstrate higher efficacy in colonic CD and UC^37–40^. Conversely, the efficacy of JAK inhibitors in colonic CD aligns with the senescence and oxidative stress pathways in *S2FCD* PDOs, which can be reversed with senolytics such as Pacritinib^7,41^. The clinical correlation of *S2FCD* with active, but mucosa-restricted colonic inflammation (mimicking UC) is likely because of stem cell dysfunction and derailed Wnt signaling that is observed in the setting of fibrosis^42^. In other words, clinical correlates observed in PDOs are not passive reflections of disease—they actively guide therapeutic alignment with disease-driving mechanisms.

Together, these data establish PDOs as NAMs capable of bridging molecular subtypes, clinical phenotypes, and therapeutic response. By abstracting essentials from patient tissues and embedding them in prospective frameworks, PDOs provide an end-to-end human-centric platform that can forecast outcomes and test therapeutic reversibility. In doing so, they transform Crohn’s disease research from descriptive biology to actionable precision medicine.

### LIMITATIONS OF STUDY

A larger cohort and longitudinal PDO assessments could enhance rigor and provide deeper insights. However, the representative nature of the cohort when it comes to the distribution of ileal vs colonic CD, and consistency of covariates support the relevance of our findings despite the sample size. Validation in an independent cohort, preferably through a prospective study, would further strengthen conclusions. Although we posit that current and emerging literature suggests the possibility that the derailment of colonic innate immune functions may trigger and/or perpetuate ileal disease, we do not define mechanisms here. Regardless of the knowledge gaps in our understanding of how regional or systemic immune differences shape ileal disease progression, our findings expose clinically meaningful correlates of fundamental cellular defects in CD that may prognosticate and inform choice of individualized, mechanism-driven therapeutic strategies in an area of unmet need.

## METHODS: SUBJECT DETAILS AND BIOINFORMATICS APPROACH

### Sex as a Biological Variable

Our study examined both male and female patients, and similar findings are reported for both sexes. Univariate analysis did reveal sex as a significant association with either molecular subtype of previously defined CD PDOs.

### Human Subjects Utilized in Current Study

Previously generated CD PDOs, which underwent comprehensive multi-omics analyses for molecular phenotyping^7^, were utilized for univariate and multivariate analyses. In brief, patients were recruited during routine colonoscopy procedures at the University of California, San Diego (UCSD) IBD Center. This study was conducted under an Institutional Review Board (IRB)-approved protocol in compliance with the Human Research Protection Program (HRPP) (Project ID# 1132632; PIs: Boland and Sandborn). Each participant provided written informed consent for the collection of colonic biopsies, the derivation of 3D organoids, and the use of associated clinical metadata for research purposes. The isolation and biobanking of organoids from colonic biopsies were conducted under a separate IRB-approved protocol (Project ID# 190105; PIs: Ghosh and Das) at the UCSD HUMANOID Center of Research Excellence (CoRE). All human subject data— including age, gender, and disease history—were de-identified and handled in accordance with HIPAA regulations. The study was designed and conducted following the ethical principles outlined in the Declaration of Helsinki.

### Clinical Data Collection & Analysis

For CD patients, univariate analyses were performed using a comprehensive set of clinical parameters, including patient demographics (i.e. gender, age, ethnicity); disease classification and phenotype, i.e. Montreal classification, history of stricturing disease, prior ileocecal resection; disease activity indices, i.e. patient reported outcomes (PRO)^43^, Simple Endoscopic Score for Crohn’s Disease (SES-CD)^44^; additional clinical parameters included therapeutics history, i.e. use of advanced therapies and treatment response, location of biopsy source, and PDO molecular subtype. See **Table S2** for full list of clinical features incorporated in univariate and multivariate linear regression analysis.

### Organoid Isolation from Colonic Specimens

PDOs were isolated from colonic tissue specimens collected from CD subjects using established crypt isolation protocols as previously summarized^7^. These intestinal crypts, which contain crypt-base columnar (CBC) cells, were enzymatically dissociated and processed for 3D organoid culture. Full details of organoid isolation, propagation, and molecular subtyping can be found in Tindle et al.^7^. Only CD subjects were included in the current study.

### Computational Methods

#### Differentially expressed genes and pathway enrichment

Differentially expressed genes were identified using the DESeq2 package in R. A volcano plot was generated using the Matplotlib library (v 3.10.1) in Python. Pathway enrichment analysis was performed using ShinyGO 0.82.

#### Univariate and multivariate regression analyses

These were conducted across all clinical parameters, with molecular subtypes treated as a binary base variable. The Ordinary Least Squares (OLS) regression was performed using the statsmodels module in Python. The p-value for each term tests the null hypothesis that the corresponding coefficient equals zero (i.e., no effect). This approach allows us to examine how molecular subtypes of colonic CD-PDOs (independent predictor variable) affects the likelihood of any clinical outcome (dependent variable), which can be either binary (e.g., yes/no, worsening/improving) or reflect a continuum of discretely scored entities (e.g., Simple Endoscopic Score [SES] of disease severity). Significant relationships between the two variables estimate the probability of the outcome based on the CD-PDO subtype.

## Data Availability

RESOURCE AVAILABILITY
Lead Contacts
Further information and requests for resources and reagents should be directed to and will be fulfilled by the lead contacts, Pradipta Ghosh, and Brigid S. Boland.
Materials availability
This study utilizes data generated from an organoid biobank. These materials are available from the lead contact with a completed Materials Transfer Agreement and patented technology agreement (Ghosh) following the guidelines of the Regents of the University of California, San Diego.
Data and code availability
Newly generated transcriptomic datasets reported in this paper involve re-analysis of GSE192819 which has been deposited in NCBI's Gene Expression Omnibus. The new GSE ID assignment is currently pending. This paper did not generate or use custom computer codes.
Any additional information required to reanalyze the data reported in this work paper is available from Lead. Contact upon request.

## LIST OF ABBREVIATIONS

AdSC: Adult stem cell–derived organoids
CD: Crohn’s disease
EIM: Extraintestinal manifestations
IBD: Inflammatory Bowel Disease
IC: Ileocecal
IDICD: Immune-Deficient Infectious CD
KEGG: Kyoto Encyclopedia of Genes and Genomes
MV: Multivariate logistic regression
non-penetrating NSNP: Non-stricturing
PDOs: Patient-derived organoids
PRO: Patient-reported outcomes
S2FCD: Stress and Senescence-Induced Fibrostenotic CD
SES-CD: Simple endoscopic score for Crohn’s disease
UC: Ulcerative colitis
UV.: Univariate logistic regression

## AUTHOR CONTRIBUTIONS

H.M.P., S.S., C.T., B.S.B., and P.G. conceptualized, supervised, and administered the project. B.S.B and P.G acquired funding to support it. Computational analyses were carried out by S.S. under the supervision of P.G. K.Z., H.N.L., J.N., and B.S.B provided key resources for human subjects and were responsible for the selection and enrollment of patients into this study. H.M.P., S.S., and P.G. prepared figures for data visualization. H.M.P. and P.G. wrote the original draft. H.M.P, S.S., C.T., B.S.B, and P.G. reviewed and edited the draft. All co-authors approved the final version of the manuscript.

## RESOURCE AVAILABILITY

### Lead Contacts

Further information and requests for resources and reagents should be directed to and will be fulfilled by the lead contacts, Pradipta Ghosh and Brigid S. Boland.

### Materials availability

This study utilizes data generated from an organoid biobank. These materials are available from the lead contact with a completed Materials Transfer Agreement and patented technology agreement (Ghosh) following the guidelines of the Regents of the University of California, San Diego.

### Data and code availability

Newly generated transcriptomic datasets of PDOs from the colons of patients with CD have been deposited in NCBI’s GEO functional genomic repository under accession number GSE293539. This paper did not generate or use custom computer codes. Any additional information required to reanalyze the data reported in this work is available from the lead contact upon request.

## SUPPLEMENTAL INFORMATION

**Table S1:** Differentially expressed genes between *IDICD* and *S2FCD* molecular subtypes of CD

**Table S2:** Excel datasheet with CD clinical parameters used for univariate and multivariate analyses in this current study

## REFERENCES

1. Baumgart, D.C., and Le Berre, C. (2021). Newer Biologic and Small-Molecule Therapies for Inflammatory Bowel Disease. N Engl J Med 385, 1302–1315. 10.1056/NEJMra1907607.

2. Torres, J., Mehandru, S., Colombel, J.F., and Peyrin-Biroulet, L. (2017). Crohn’s disease. Lancet 389, 1741–1755. 10.1016/S0140-6736(16)31711-1.

3. Torres, J., and Colombel, J.F. (2016). Genetics and phenotypes in inflammatory bowel disease. Lancet 387, 98–100. 10.1016/S0140-6736(15)00464-X.

4. DeVoss, J., and Diehl, L. (2014). Murine models of inflammatory bowel disease (IBD): challenges of modeling human disease. Toxicol Pathol 42, 99–110. 10.1177/0192623313509729.

5. Lee, C., An, M., Joung, J.G., Park, W.Y., Chang, D.K., Kim, Y.H., and Hong, S.N. (2022). TNFalpha Induces LGR5+ Stem Cell Dysfunction In Patients With Crohn’s Disease. Cell Mol Gastroenterol Hepatol 13, 789–808. 10.1016/j.jcmgh.2021.10.010.

6. d’Aldebert, E., Quaranta, M., Sebert, M., Bonnet, D., Kirzin, S., Portier, G., Duffas, J.P., Chabot, S., Lluel, P., Allart, S., et al. (2020). Characterization of Human Colon Organoids From Inflammatory Bowel Disease Patients. Front Cell Dev Biol 8, 363. 10.3389/fcell.2020.00363.

7. Tindle, C., Fonseca, A.G., Taheri, S., Katkar, G.D., Lee, J., Maity, P., Sayed, I.M., Ibeawuchi, S.R., Vidales, E., Pranadinata, R.F., et al. (2024). A living organoid biobank of patients with Crohn’s disease reveals molecular subtypes for personalized therapeutics. Cell Rep Med 5, 101748. 10.1016/j.xcrm.2024.101748.

8. Mills, S., and Stamos, M.J. (2007). Colonic Crohn’s disease. Clin Colon Rectal Surg 20, 309–313. 10.1055/s-2007-991030.

9. Huch, M., Knoblich, J.A., Lutolf, M.P., and Martinez-Arias, A. (2017). The hope and the hype of organoid research. Development 144, 938–941. 10.1242/dev.150201.

10. Poetsch, M.S., Strano, A., and Guan, K. (2022). Human Induced Pluripotent Stem Cells: From Cell Origin, Genomic Stability, and Epigenetic Memory to Translational Medicine. Stem Cells 40, 546–555. 10.1093/stmcls/sxac020.

11. Doss, M.X., and Sachinidis, A. (2019). Current Challenges of iPSC-Based Disease Modeling and Therapeutic Implications. Cells 8. 10.3390/cells8050403.

12. Zhao, D., Ravikumar, V., Leach, T.J., Kraushaar, D., Lauder, E., Li, L., Sun, Y., Oravecz-Wilson, K., Keller, E.T., Chen, F., et al. (2024). Inflammation-induced epigenetic imprinting regulates intestinal stem cells. Cell Stem Cell 31, 1447–1464 e1446. 10.1016/j.stem.2024.08.006.

13. van de Wetering, M., Francies, H.E., Francis, J.M., Bounova, G., Iorio, F., Pronk, A., van Houdt, W., van Gorp, J., Taylor-Weiner, A., Kester, L., et al. (2015). Prospective derivation of a living organoid biobank of colorectal cancer patients. Cell 161, 933–945. 10.1016/j.cell.2015.03.053.

14. Dennison, T.W., Edgar, R.D., Payne, F., Nayak, K.M., Ross, A.D.B., Cenier, A., Glemas, C., Giachero, F., Foster, A.R., Harris, R., et al. (2024). Patient-derived organoid biobank identifies epigenetic dysregulation of intestinal epithelial MHC-I as a novel mechanism in severe Crohn’s Disease. Gut 73, 1464–1477. 10.1136/gutjnl-2024-332043.

15. Geng, T., Bao, S., Sun, X., Ma, D., Zhang, H., Ge, Q., Liu, X., and Ma, T. (2023). A clarification of concepts related to the digestion and absorption of carotenoids and a new standardized carotenoids bioavailability evaluation system. Food Chem 400, 134060. 10.1016/j.foodchem.2022.134060.

16. Geyer, J., Wilke, T., and Petzinger, E. (2006). The solute carrier family SLC10: more than a family of bile acid transporters regarding function and phylogenetic relationships. Naunyn Schmiedebergs Arch Pharmacol 372, 413–431. 10.1007/s00210-006-0043-8.

17. Richter, A.N., and Khoshgoftaar, T.M. (2018). A review of statistical and machine learning methods for modeling cancer risk using structured clinical data. Artif Intell Med 90, 1–14. 10.1016/j.artmed.2018.06.002.

18. Yao, Y., Xu, X., Yang, L., Zhu, J., Wan, J., Shen, L., Xia, F., Fu, G., Deng, Y., Pan, M., et al. (2020). Patient-Derived Organoids Predict Chemoradiation Responses of Locally Advanced Rectal Cancer. Cell Stem Cell 26, 17–26 e16. 10.1016/j.stem.2019.10.010.

19. Ooft, S.N., Weeber, F., Dijkstra, K.K., McLean, C.M., Kaing, S., van Werkhoven, E., Schipper, L., Hoes, L., Vis, D.J., van de Haar, J., et al. (2019). Patient-derived organoids can predict response to chemotherapy in metastatic colorectal cancer patients. Sci Transl Med 11. 10.1126/scitranslmed.aay2574.

20. Vlachogiannis, G., Hedayat, S., Vatsiou, A., Jamin, Y., Fernandez-Mateos, J., Khan, K., Lampis, A., Eason, K., Huntingford, I., Burke, R., et al. (2018). Patient-derived organoids model treatment response of metastatic gastrointestinal cancers. Science 359, 920–926. 10.1126/science.aao2774.

21. Wang, T., Tang, Y., Pan, W., Yan, B., Hao, Y., Zeng, Y., Chen, Z., Lan, J., Zhao, S., Deng, C., et al. (2023). Patient-Derived Tumor Organoids Can Predict the Progression-Free Survival of Patients With Stage IV Colorectal Cancer After Surgery. Dis Colon Rectum 66, 733–743. 10.1097/DCR.0000000000002511.

22. Tiriac, H., Belleau, P., Engle, D.D., Plenker, D., Deschenes, A., Somerville, T.D.D., Froeling, F.E.M., Burkhart, R.A., Denroche, R.E., Jang, G.H., et al. (2018). Organoid Profiling Identifies Common Responders to Chemotherapy in Pancreatic Cancer. Cancer Discov 8, 1112–1129. 10.1158/2159-8290.CD-18-0349.

23. Wensink, G.E., Elias, S.G., Mullenders, J., Koopman, M., Boj, S.F., Kranenburg, O.W., and Roodhart, J.M.L. (2021). Patient-derived organoids as a predictive biomarker for treatment response in cancer patients. NPJ Precis Oncol 5, 30. 10.1038/s41698-021-00168-1.

24. Weiser, M., Simon, J.M., Kochar, B., Tovar, A., Israel, J.W., Robinson, A., Gipson, G.R., Schaner, M.S., Herfarth, H.H., Sartor, R.B., et al. (2018). Molecular classification of Crohn’s disease reveals two clinically relevant subtypes. Gut 67, 36–42. 10.1136/gutjnl-2016-312518.

25. T, N., L, R., A, K., R, P., I, A., FM, K., and U, G. (2016 Feb). Distinct Microbiotas are Associated with Ileum-Restricted and Colon-Involving Crohn’s Disease - PubMed. Inflammatory bowel diseases 22. 10.1097/MIB.0000000000000662.

26. Gonzalez, C.G., Mills, R.H., Zhu, Q., Sauceda, C., Knight, R., Dulai, P.S., and Gonzalez, D.J. (2022). Location-specific signatures of Crohn’s disease at a multi-omics scale. Microbiome 10, 133. 10.1186/s40168-022-01331-x.

27. Kong, L., Pokatayev, V., Lefkovith, A., Carter, G.T., Creasey, E.A., Krishna, C., Subramanian, S., Kochar, B., Ashenberg, O., Lau, H., et al. (2023). The landscape of immune dysregulation in Crohn’s disease revealed through single-cell transcriptomic profiling in the ileum and colon. Immunity 56, 444–458 e445. 10.1016/j.immuni.2023.01.002.

28. Kredel, L.I., Jodicke, L.J., Scheffold, A., Grone, J., Glauben, R., Erben, U., Kuhl, A.A., and Siegmund, B. (2019). T-cell Composition in Ileal and Colonic Creeping Fat - Separating Ileal from Colonic Crohn’s Disease. J Crohns Colitis 13, 79–91. 10.1093/ecco-jcc/jjy146.

29. Cleynen, I., Boucher, G., Jostins, L., Schumm, L.P., Zeissig, S., Ahmad, T., Andersen, V., Andrews, J.M., Annese, V., Brand, S., et al. (2016). Inherited determinants of Crohn’s disease and ulcerative colitis phenotypes: a genetic association study. Lancet 387, 156–167. 10.1016/S0140-6736(15)00465-1.

30. Parkes, G.C., Whelan, K., and Lindsay, J.O. (2014). Smoking in inflammatory bowel disease: impact on disease course and insights into the aetiology of its effect. J Crohns Colitis 8, 717–725. 10.1016/j.crohns.2014.02.002.

31. Subramanian, S., Ekbom, A., and Rhodes, J.M. (2017). Recent advances in clinical practice: a systematic review of isolated colonic Crohn’s disease: the third IBD? Gut 66, 362–381. 10.1136/gutjnl-2016-312673.

32. Villafruela Cives, M. (2009). Re: “The risk of oral contraceptives in the etiology of inflammatory bowel disease: a meta-analysis”. Response to Cornish et al. Am J Gastroenterol 104, 534–535. 10.1038/ajg.2008.70.

33. Richard, N., Savoye, G., Leboutte, M., Amamou, A., Ghosh, S., and Marion-Letellier, R. (2023). Crohn’s disease: Why the ileum? World J Gastroenterol 29, 3222–3240. 10.3748/wjg.v29.i21.3222.

34. Rieder, F., Zimmermann, E.M., Remzi, F.H., and Sandborn, W.J. (2013). Crohn’s disease complicated by strictures: a systematic review. Gut 62, 1072–1084. 10.1136/gutjnl-2012-304353.

35. Sahoo, D., Swanson, L., Sayed, I.M., Katkar, G.D., Ibeawuchi, S.R., Mittal, Y., Pranadinata, R.F., Tindle, C., Fuller, M., Stec, D.L., et al. (2021). Artificial intelligence guided discovery of a barrier-protective therapy in inflammatory bowel disease. Nat Commun 12, 4246. 10.1038/s41467-021-24470-5.

36. Katkar, G.D., Sayed, I.M., Anandachar, M.S., Castillo, V., Vidales, E., Toobian, D., Usmani, F., Sawires, J.R., Leriche, G., Yang, J., et al. (2022). Artificial intelligence-rationalized balanced PPARalpha/gamma dual agonism resets dysregulated macrophage processes in inflammatory bowel disease. Commun Biol 5, 231. 10.1038/s42003-022-03168-4.

37. Atreya, R., Bojarski, C., Kuhl, A.A., Trajanoski, Z., Neurath, M.F., and Siegmund, B. (2022). Ileal and colonic Crohn’s disease: Does location makes a difference in therapy efficacy? Curr Res Pharmacol Drug Discov 3, 100097. 10.1016/j.crphar.2022.100097.

38. Arnott, I.D., McNeill, G., and Satsangi, J. (2003). An analysis of factors influencing short-term and sustained response to infliximab treatment for Crohn’s disease. Aliment Pharmacol Ther 17, 1451–1457. 10.1046/j.1365-2036.2003.01574.x.

39. Dulai, P.S., Boland, B.S., Singh, S., Chaudrey, K., Koliani-Pace, J.L., Kochhar, G., Parikh, M.P., Shmidt, E., Hartke, J., Chilukuri, P., et al. (2018). Development and Validation of a Scoring System to Predict Outcomes of Vedolizumab Treatment in Patients With Crohn’s Disease. Gastroenterology 155, 687–695 e610. 10.1053/j.gastro.2018.05.039.

40. Lee, H.H., Yuan, Y., Boland, B.S., Chang, J.T., Jairath, V., and Singh, S. (2025). Efficacy of Advanced Therapies in Achieving Remission by Disease Location in Crohn’s Disease: A Systematic Review and Meta-analysis. Clin Gastroenterol Hepatol. 10.1016/j.cgh.2025.07.009.

41. Xu, M., Tchkonia, T., Ding, H., Ogrodnik, M., Lubbers, E.R., Pirtskhalava, T., White, T.A., Johnson, K.O., Stout, M.B., Mezera, V., et al. (2015). JAK inhibition alleviates the cellular senescence-associated secretory phenotype and frailty in old age. Proc Natl Acad Sci U S A 112, E6301–6310. 10.1073/pnas.1515386112.

42. Kinchen, J., Chen, H.H., Parikh, K., Antanaviciute, A., Jagielowicz, M., Fawkner-Corbett, D., Ashley, N., Cubitt, L., Mellado-Gomez, E., Attar, M., et al. (2018). Structural Remodeling of the Human Colonic Mesenchyme in Inflammatory Bowel Disease. Cell 175, 372–386 e317. 10.1016/j.cell.2018.08.067.

43. Williet, N., Sandborn, W.J., and Peyrin-Biroulet, L. (2014). Patient-reported outcomes as primary end points in clinical trials of inflammatory bowel disease. Clin Gastroenterol Hepatol 12, 1246–1256 e1246. 10.1016/j.cgh.2014.02.016.

44. Daperno, M., D’Haens, G., Van Assche, G., Baert, F., Bulois, P., Maunoury, V., Sostegni, R., Rocca, R., Pera, A., Gevers, A., et al. (2004). Development and validation of a new, simplified endoscopic activity score for Crohn’s disease: the SES-CD. Gastrointest Endosc 60, 505–512. 10.1016/s0016-5107(04)01878-4.

